# When do we need massive computations to perform detailed COVID-19 simulations?

**DOI:** 10.1101/2021.08.26.21262694

**Authors:** Christopher B. Lutz, Philippe J. Giabbanelli

## Abstract

The COVID-19 pandemic has infected over 200 million people worldwide and killed more than 4 million as of August 2021. Many intervention strategies have been utilized by governments around the world, including masks, social distancing, and vaccinations. However, officials making decisions regarding interventions may have a limited time to act. Computer simulations can aid them by predicting future disease outcomes, but they also have limitations due to requirements on processing power or time. This paper examines whether a machine learning model can be trained on a small subset of simulation runs to inexpensively predict future disease trajectories very close to the original simulation results. Using four previously published agent-based models for COVID-19, this paper analyzes the predictions of decision tree regression machine learning models and compares them to the results of the original simulations. The results indicate that accurate machine learning meta-models can be generated from simulation models with no strong interventions (e.g., vaccines, lockdowns) using small amounts of simulation data. However, meta-models for simulation models that include strong interventions required much more training data to achieve a similar accuracy. This indicates that machine learning meta-models could be used in some scenarios to assist in faster decision making.

## 1 Introduction

As of August 2021, COVID-19 was directly responsible for an estimate of over 4 million deaths and over 200 million cases ^[1]^. Taking the USA as an example, these numbers translate to almost 650,000 deaths from about 40 million cases ^[2]^, while noting that fatality may be under-estimated ^[3]^ and differs across sub-groups based on factors such as socio-economic status or race and ethnicity ^[4]^. Although reports previously considered that “transition toward normalcy in the United States remains most likely in the second quarter of 2021” ^[5]^, the delta variant has effectively triggered a ‘new phase in the pandemic’ ^[6]^ as can be seen with a rebound of over 100,000 new cases per day in the USA ^[2]^. Similar phenomena can be observed worldwide and continue to require action by government officials to limit the spread of disease ^[7]^. For example, France has implemented a COVID-19 ‘health pass’ while Italy has a similar ‘Green Pass’; mandates for masks or vaccines are on the agenda across several US states; and lockdown as well as travel curbs are making a come-back in China. A broad set of intervention strategies is available to policymakers ^[8, 9, 10, 11]^, including vaccines, preventative care (e.g., masks and social distancing), or lockdowns (e.g., remote work and travel restrictions). Implementing one of these strategies involves several logistical parameters: for instance, testing requires capacity for contact tracing and policies for quarantine (e.g., is a negative test required to leave quarantine? is a number of days required?); similarly, vaccination involves a complex logistical chain from shipping to administering doses ^[12]^.

In addition to the many possible combinations of interventions and parameter values, the ‘right set’ of interventions can vary across places (e.g., based on disease incidence and hospital capacity), across time (e.g., in response to a new wave of infections), and across individuals (e.g., priority for vaccination to those most at risk). This results in a *very large search space of possible interventions* ^[13]^. Although the necessity of immediate actions in the early days of the pandemic may have resulted in choosing an intervention based on minimal insight, there is now evidence for serious consequences in rolling a sub-optimal intervention: lives may be lost, the cost of future interventions may be heightened, or the adherence (hence the impact) of future interventions may be lowered ^[14, 15, 16]^.

Computer simulations using *Agent-Based Models* (ABMs) can aid officials in making these decisions, by modeling the effects of specific interventions in specific places (e.g., small towns ^[17]^, educational institutions ^[18, 19]^, supermarkets ^[20]^), populations (e.g., targeted vaccinations ^[21]^), or time windows (e.g., during a yearly mass pilgrimage ^[22]^). Several modeling frameworks are available to quickly run simulations for specific interventions, while accounting for individual heterogeneity in risk factors and contact patterns (e.g., by embedding agents across multiple networks such as community and work). However, *significant computing resources are required* to perform detailed simulations that track individual transmissions and evaluate various interventions ^[23, 24]^. Our analysis across six projects showed that cloud computing or high performance computing clusters were frequently needed ^[25]^. This is exemplified by the model from Chang *et al*., which ran on 4,264 compute cores ^[23]^.

So far, the primary solution to perform resource-intensive agent-based COVID-19 simulations has been to find more computing power. For example, the COVID-19 High-Performance Computing Consortium (covid19-hpc-consortium.org) was created to make private computing resources available to COVID-19 researchers ^[26]^. Similarly, the Partnership for Advanced Computing in Europe (PRACE) offered a fast-track mechanism for access to supercomputers (prace-ri.eu/hpc-access/hpcvsvirus), and national laboratories issues calls for rapid-response research ^[27]^. These computing requirements can be limiting in the context of pandemic responses, because officials are forced to wait for results before acting to prevent the spread of the disease. It also stresses inequities in simulation research, as some groups may struggle to perform their simulations in a timely manner due to the lack of resources at smaller research organizations ^[28]^.

In this paper, we examine whether massive computations are an absolute requirement to support decision-makers in comprehensively examining the expected consequences of various intervention scenarios in the context of COVID-19. Specifically, we assess whether a machine learning model can be trained on *few* expensive simulations and predict the remaining results inexpensively. In other words, we evaluate the feasibility of using machine learning models as computationally inexpensive proxies (or *meta-models*) to expensive COVID-19 simulations. Our approach consists of generating data from four previously published and validated agent-based models (ABMs) for COVID-19 ^[12, 24, 29, 30]^ and using the data to train machine learning regression meta-models. By varying the amount of data used to train these meta-models, we characterize the relationship between how much data is used to train the meta-model and how accurate that meta-model is. Our specific contributions are as follows:

- We develop machine learning regression meta-models for four COVID-19 agent-based models to predict the total proportion of the population that will become infected, in response to the intervention scenarios captured by the model’s parameters.
- We examine the affects that different amounts of training data have on the overall accuracy of those meta-models, thus establishing the situations under which a COVID-19 ABM may require computing power to achieve accurate results.

The remainder of this paper is organized as follows. In section 2, we summarize the key features of COVID-19 and interventions that are available in the four ABMs used in this study. Our background also briefly explains how agent-based models are created for COVID-19 and how machine learning can be used to generate simulation meta-models. Then in section 3, we cover our methods starting with the implementation and verification of the selected agent-based models, and then detailing how we performed our machine learning regressions. In section 4, we analyze the results produced by the verification data for our agent-based models and the machine learning regressions. Next in section 5, we discuss the interpretation and limitations of our results. Finally in section 6, we provide concluding remarks and suggest future work that could be undertaken based on our results.

## 2 Background

In this section, we will examine key details of COVID-19 and available interventions, as well as how agent-based models are created for it. Then, we summarize how machine learning has been used previously to create meta-models for simulations.

### 2.1 COVID-19

COVID-19, caused by the SARS-CoV-2 virus, was first reported in Wuhan, China in December, 2019 ^[31]^ with newer studies suggesting a first case as far back as mid-November, 2019 ^[32]^. The virus spreads through droplets that are released when an infected individual coughs or sneezes. As a respiratory disease, the primary mode of transmission is thus via exposure to droplets, either indirectly (e.g., via contact through contaminated objects or hands) or directly (air borne) ^[33, 34]^. The virus infects cells in an individual’s lungs, interfering with the lungs’ ability to function properly ^[35]^. Symptoms for the disease include fever, loss of smell, or cough ^[36, 37]^. A systematic review of 45 studies reported that 73% of individuals experience at least one persistent symptom ^[38]^. Most commonly, symptoms such as fatigue, sleep disorders, or loss of smell can be experienced for months ^[39]^. Less common consequences include multi-organ damage ^[40]^, for example in the the cardiovascular system ^[41]^, kidneys ^[42]^, nervous system ^[43]^, or immune system ^[44]^.

Prior to the development of vaccines, all interventions for COVID-19 were necessarily *non-pharmaceutical*. These interventions included the use of masks, social distancing, regional lockdowns, and contact tracing. Masks reduce the spread of COVID-19 by lowering the potential for infected particles from entering the environment, and higher mask compliance leads to more effective disease mitigation ^[45]^. Social distancing and the closing of restaurants, gyms, and other public locations led to a statistically significant reduction in the spread of COVID-19 as well ^[46]^. Contact tracing, which helps officials control the spread of the virus by tracking who may have been in contact with an infected individual, has also been found to be effective ^[47]^.

In December of 2020, the first *pharmaceutical* intervention (i.e. vaccine) for COVID-19 became available ^[48]^. The messenger RNA (mRNA) vaccines for COVID-19 contain a small piece of the SARS-CoV-2 virus’s mRNA that triggers the immune system to start producing an immune response to the virus. The FDA found that the Pfizer vaccine was 95% effective against COVID-19 ^[48]^, but effectiveness may be reduced by mutations ^[49]^. Mutations known as ‘variants of concern’ impact aspects such as transmissibility and vaccine effectiveness ^[50]^. In particular, some variants can evade antibodies caused by infection and current vaccines ^[51]^, which calls for the development of next-generation vaccines and antibody therapies ^[52]^. The existence of repeat infections ^[53]^ as well as breakthrough infections ^[54, 55]^ (i.e., infection of a fully vaccinated person) have led to question the possibility of herd immunity ^[56]^, that is, the assumption that most transmission would be blocked if a given threshold of the population gains immunity.

### 2.2 Agent-Based Models For COVID-19

In early 2020, given the urgency of the COVID-19 pandemic and the scarcity of data, the first generation of COVID-19 models was based on a compartmental approach in which individuals are aggregated into groups (e.g., susceptible, infected) and a simulation proceeds by applying flow equations between groups. These classic compartmental epidemiological models focused on estimating key epidemiological quantities such as *R*_0_, the number of new cases generated by each infected individual ^[57, 58]^. Although these models were imperfect in different ways ^[59, 60, 61]^, expert predictions have still outperformed the general public ^[62]^ and their models “have influenced public health policies and increased the familiarity of the general public as well as policy-makers with the modeling process, its value, and its limitations” ^[25]^. The most commonly used epidemic models categorized the population into susceptible, infected, and recovered (SIR) or added an intermediate stage for exposure (SEIR) ^[63]^. A review of models produced between January and November 2020 found that the SIR and SEIR approaches represented 46.1% of all models, whereas Agent-Based Models (ABM) only accounted for 1.3% of studies at the time ^[64]^. As the evidence-base progressed, modelers realized that the assumptions of compartmental models (e.g., treating individuals as part of homogeneous groups) were limiting. In the words of Tolk and colleagues, “As the pandemic unfolded, it quickly became evident these were not valid assumptions: the virus does not impact all populations evenly, and the interaction among different groups is far from even.” ^[65]^ The rationale for the use of individual-level simulation models such as ABM thus centers on the notion of *heterogeneity* : heterogeneity of risk factors for individuals (e.g., age and underlying or ‘pre-existing’ conditions that increase the risk of severe illness conditions ^[66, 67]^), heterogeneity of behaviors (e.g., interest in vaccination ^[68]^, compliance with mask policies ^[69, 70]^), heterogeneity in socio-ecological vulnerability across places (e.g., lower access to resources in rural counties ^[71]^, urban sub-populations at risk ^[72]^), and heterogeneity in contact patterns ^[73, 74]^. Although the need was clear, the development of ABMs was initially challenged by a lack of data, limited understanding of the disease, and occasionally a limited skillset ^[25]^. The situation has changed with the growing evidence-base on COVID-19, the availability of mobility data, and the development of reusable frameworks to instantiate ABMs specifically for COVID-19 (e.g., COVASIM ^[75]^, OpenABM-Covid19 ^[76]^, various modeling pipelines ^[77]^).

An Agent-Based model represents individuals (as agents) and their interactions with the environment as well as other agents. Each agent can have a state along with additional characteristics such as age or position within social networks. States in several ABMs are inspired by classic compartmental epidemiological models, hence it is common to use a SEIR model to represent the progression of each agent ^[29, 78, 79]^. Although earlier models may have relied on only four stages (SEIR), newer ones have introduced new stages to account for vaccination in two doses, disease severity, and the difference between asympatomatic and symptomatic individuals (**Figure 1**). In contrast with their mathematical predecessors, ABMs include the movements of agents over a space (e.g., a grid) and interactions with other agents or their environment (e.g., contamined surfaces), which leads to the spread of the simulated disease. One of the factors that makes ABM agents unique is their ability to model individual characteristics such as age or other risk factors in the context of infectious disease.

**Figure 1:**
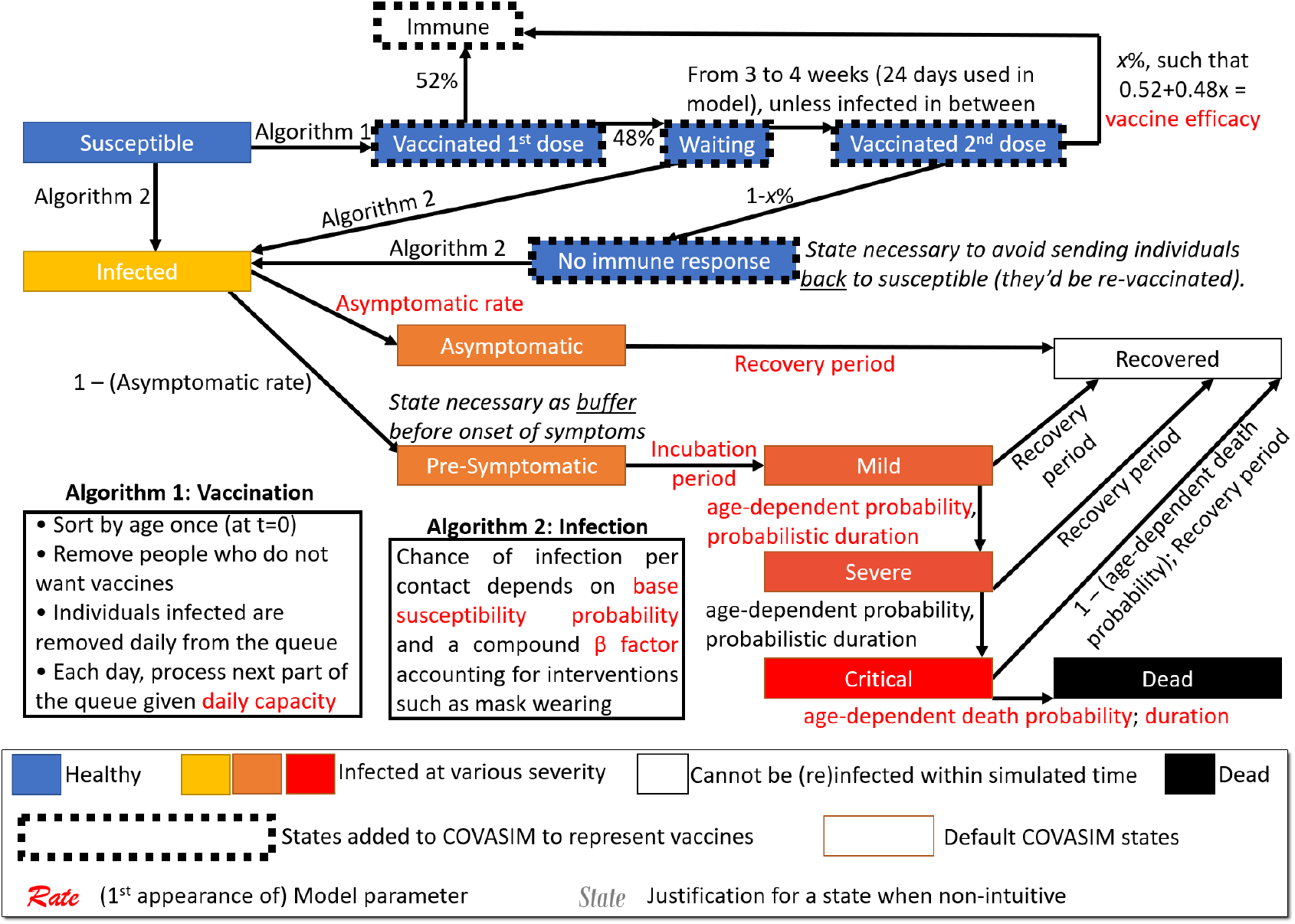
Flow diagram describing the possible journey of an agent, in the study of Li and Giabbanelli ^[12]^. Additional model logic is governed by algorithms, to identify agents who will be vaccinated or infected.

States and characteristics found in the four chosen models for our study are expanded in **Table 1**. Although each model has been previously published with a detailed technical description, we provide a succinct overview of each model in the subsequent subsections to keep this manuscript self-contained. Since a key motivation for our approach (and the use of meta-modeling in general) is to create a *cheaper* proxy to a computationally expensive model, we exemplify the wallclock time typically required by the four chosen models, across platforms (**Table 2**). These platforms show the diversity of hardware that end-users of models can access, from cloud-computing services (Microsoft Azure with AMD EPYC platform for Li and Giabbanelli ^[12]^) and High-Performance Computing clusters (Intel Xeon processors with 96 Gb of memory for Shamil *et al*. ^[24]^ and Silva *et al*. ^[29]^) to personal computers (AMD Ryzen with 16 Gb of memory for Badham *et al*. ^[30]^).

**Table 1:** Characteristics of the agent-based models.

| Characteristic | Li <sup>[12]</sup> | Shamil <sup>[24]</sup> | Silva <sup>[29]</sup> | Badham <sup>[30]</sup> |
| --- | --- | --- | --- | --- |
| **Model States** |  |  |  |  |
| Healthy (Susceptible) | Yes | Yes | Yes | Yes |
| Infected (Not Contagious) | Yes | Yes |  | Yes |
| Infected (Contagious, Asymptomatic) | Yes | Yes | Yes |  |
| Infected (Contagious, Symptomatic) | Yes | Yes | Yes | Yes |
| Hospitalized |  |  | Yes | Yes |
| Mild Infection | Yes |  |  |  |
| Severe Infection | Yes |  | Yes |  |
| Critical Infection | Yes |  |  | Yes |
| Recovered (Immune) | Yes | Yes | Yes | Yes |
| Dead | Yes | Yes | Yes | Yes |
| Vaccinated (First Dose) | Yes |  |  |  |
| Vaccinated (Waiting) | Yes |  |  |  |
| Vaccinated (Second Dose) | Yes |  |  |  |
| Fully Vaccinated | Yes |  |  |  |
| **Model Characteristics** |  |  |  |  |
| Time Resolution | Days | Hours | Hours | Days |
| Work Network | Yes | Yes | Yes |  |
| School Network | Yes | Yes |  |  |
| Community Network | Yes | Yes | Yes | Yes |
| Home Network | Yes | Yes | Yes |  |
| Number of Agents | 656,000 | 10,000 | Varies | 20,172 |

**Table 2:** Examples of wallclock time across hardware configurations for the four COVID-19 ABMs considered here.

| Model | Wallclock time per simulation run (seconds) | Hardware setup |
| --- | --- | --- |
| Li <sup>[12]</sup> | Between 900 and 1,200 | Cloud computing (Azure) |
| Shamil <sup>[24]</sup> | 32,492.86 on average | High Performance Computing cluster |
| Silva <sup>[29]</sup> | 727.12 on average | High Performance Computing cluster |
| Badham <sup>[30]</sup> | 34.63 on average | Personal computer |

#### 2.2.1 Li and Giabbanelli Model ^[12]^

The ABM developed by Li and Giabbanelli ^[12]^ is built on the Covasim framework, by including several modifications that introduce vaccination. This ABM uses 656,000 agents and simulates the spread of COVID-19 for 180 days. This model contains states for susceptible agents, as well as several different states of infection, including asymptomatic infection, and three other levels of infection from mild to critical (**Figure 1**). It also includes the ability to simulate two vaccine doses for agents, which can remove them from the pool of susceptible agents. In addition to vaccine support, the model also inherits the ability to handle quarantines and contact tracing from Covasim. Finally, the Covasim platform embeds agents across different networks (e.g., community, work, school) to model how interventions have different impacts across settings (e.g., social distancing in the community, face masks at work).

#### 2.2.2 Shamil et al. Model ^[24]^

The ABM created by Shamil *et al*. ^[24]^ includes two possible agent configurations for different cities in the United States. One such configuration is a simplified representation for New York City, which contains 10,000 agents simulated for 90 days. It contains states representing different states of contagion and symptoms, with agents initially healthy and transitioning through a non-contagious state into a contagious asymptomatic state, then into a contagious symptomatic state, and finally into a recovered or dead state. This model includes representations of large-scale gatherings and various other day-to-day activities that could spread the virus, such as attending work or school. This ABM provides support for contact tracing and quarantines.

#### 2.2.3 Silva et al. Model ^[29]^

The ABM released by Silva *et al*. ^[29]^ models agents in infected states, as well as different levels of virus severity and hospitalization. Age and risk factors are also included for each agent, and these can be used when determining a lockdown policy. This simulation models the interactions of different agents from different home environments as well, and allows for lockdowns that prevent agents from traveling and interacting. This ABM provides support for quarantines, lockdowns, and masks.

#### 2.2.4 Badham et al. Model ^[30]^

The ABM built in Badham *et al*. ^[30]^ models agents in infected states, and contains states for hospitalization and critical infection. Community spread is represented as the agents move around their environment, although movement of agents can be restricted by several policies. This ABM provides support for social distancing, hospital simulation, isolation, and movement restrictions.

### 2.3 Simulation Meta-Modeling

Meta-models, also known as surrogate models, are *approximations* of a more complex model ^[80]^. Although an approximation may be less accurate, this is usually tolerated in exchange for a significant improvement (i.e., reduction) in computational cost such as wallclock time ^[80]^. For example, a meta-model for hydrodynamic and thermal simulation reduced compute time by 93% while only reducing accuracy by 4% ^[81]^. In another simulation of social networks, simplifications resulted in an 85% decrease in runtime and a 32% decrease in memory requirements ^[82]^.

As the notion of ‘models’ can be broadly conceptualized across fields, it is important to distinguish two settings. In pure mathematics, meta-models are mathematical functions that approximate the output of another, more complex mathematical function. In the simulation of interest here, meta-models are models that predict the results of a simulation with less computational requirements. Machine learning is one approach to create simulation meta-models. One common type of machine learning meta-model is a regression model ^[83][84][85]^, which is appropriate when the output of the simulation model (which we seek to approximate) is a discrete value based on its input parameters. Another common type of machine learning meta-model is a classification model, which produces produce a *class* (i.e. a group with similar characteristics) based on input parameters ^[86]^. In this paper, we focus on regressions, whereby the objective is to provide a accurate but faster proxy to the *final* result of an expensive simulation.

**Figure 2** provides a high-level illustration for the process for constructing a simulation meta-model via machine learning. Gathering data for a meta-model involves running the simulation multiple times with varying input parameters. The parameters that should be varied during simulation runs are the desired meta-model input parameters ^[87]^. For example, to train a clinically-relevant meta-model of the Human Immunodeficiency Virus (HIV), a HIV simulation model may be run for several values of clinically-relevant parameters such as the start and expected efficacy of treatment ^[88]^. The data produced by the simulation model then becomes the input or training data for the meta-model. The number of data points gathered for training depends on the computational requirements of the simulation itself ^[89]^. Once the meta-model has been trained, its results are compared against the simulation itself using an error metric such as the mean-square error (MSE), root-mean-square error (RMSE) ^[87]^, or normalized root mean square error ^[90]^.

**Figure 2:**
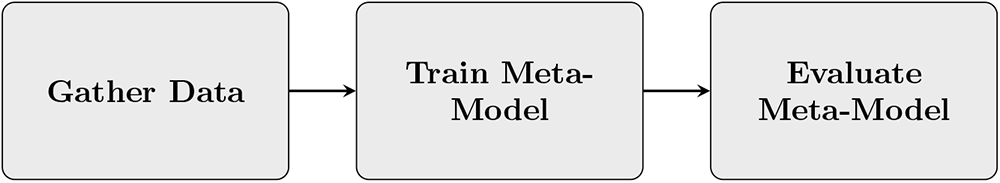
Flowchart for the process of training a meta-model.

Several sampling methods or ‘designed experiments’ allow to produce the training data for the metamodel, that is, select values of the simulation parameters. Such techniques include random sampling, factorial sampling, and Latin hypercube sampling. Random sampling picks a certain number of entries or parameter values at random, which may lead to a cluster of points (i.e., over-sampling in some areas) or an absence of points (i.e., under-sampling). Factorial sampling explores every value of a parameter combined with every value of every other parameter ^[91]^. For example, if two different parameters could be either 0 or 1, the factorial set would be (0, 0), (0, 1), (1, 0), and (1, 1). Latin hypercube sampling uses each value for a parameter only once, but ensures an even spread over the domain of parameter values ^[92]^.

## 3 Methods

In this section, we explain how we used each agent-based models to perform the machine learning regressions. A table is provided to list the parameter values used in generating data from each model. When the parameter values require further explanations, a secondary table is also included; additional details can be found in the peer-reviewed manuscripts corresponding to each model. Apart from the Li and Giabbanelli model for which we already had data (as it was produced by our group), we accessed the public implementations for each of their other models, verified the code vis-a-vis the corresponding publication, and then produced the simulation data. A flowchart of the overall process is provided in **Figure 3**.

**Figure 3:**
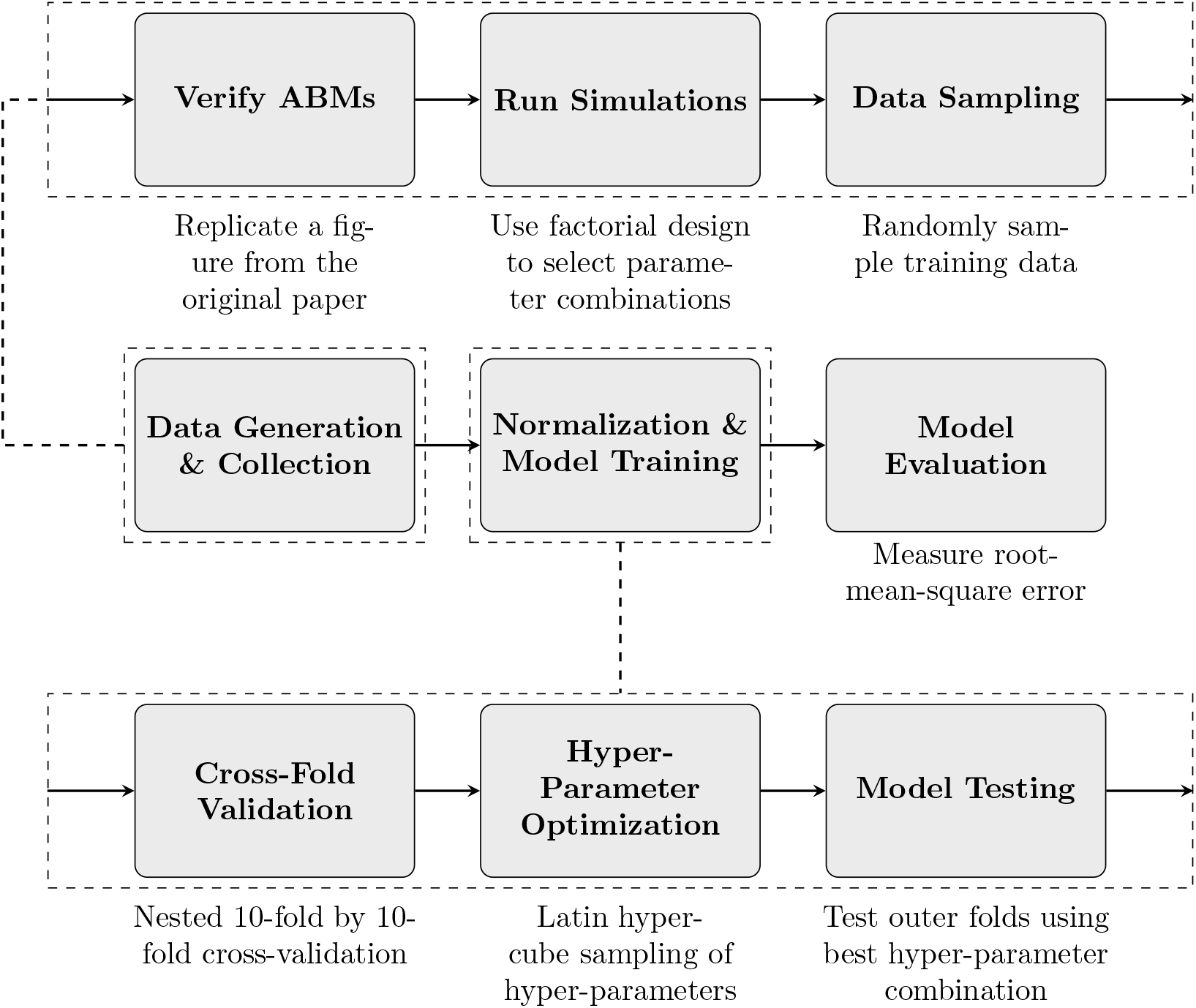
Our process for training a machine learning regression meta-model, detailing data generation from simulations (top) and machine learning (bottom).

### 3.1 Data Generation & Collection

The datasets generated based on the following procedures can all be openly accessed by readers from our third-party repository, https://osf.io/d7vqa/.

#### 3.1.1 Li and Giabbanelli Model ^[12]^

This model was produced by our research group and utilized as part of a factorial analysis (in the absence of vaccines) ^[13]^ or via a grid search (with vaccines) ^[12]^. It was thus the only case in which we had an existing and extensive simulated dataset to rely upon. For our meta-modeling purposes, we split the original dataset into three separate sets based on their vaccine plans: no vaccines in **Table 4** and the two vaccination plans in **Table 3**. The first subset of data captured simulations that did not include a vaccine, the second subset contained the simulations that used one vaccine plan (under the Trump administration), and the third subset contained was formed of vaccine simulations under the other vaccine plan (from the current Biden administration). These three sets of data were used to train three different meta-models, since the two vaccine datasets contained vaccination while the first, non-vaccine dataset was limited to non-pharmaceutical interventions. The number of simulation runs per combination of parameter values varies, as it was set to automatically spawn new runs until a 95% confidence interval would be reached (in contrast with a pre-determined *fixed* number of runs). The dataset had a total of *n* = 727, 706 data points, which are approximately evenly divided between the vaccine subsets (*n* = 209, 042 for the Trump plan and *n* = 207, 990 for the Biden plan) and the non-pharmaceutical subset (*n* = 310, 674).

**Table 3:**
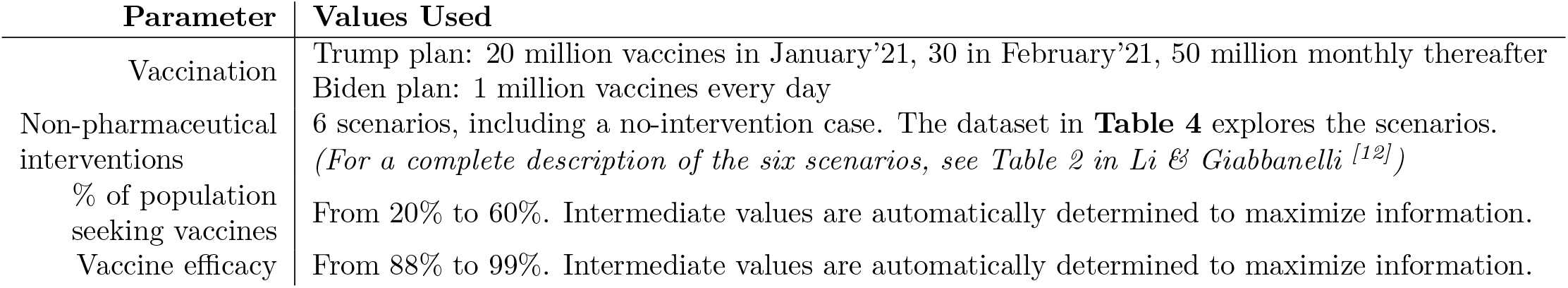
Parameters and values used by Li and Giabbanelli. The two intervention settings (Biden, Trump) are run separately. Each intervention setting is run with all combination of the other parameter values.

| Parameter | Values Used |
| --- | --- |
| Vaccination | Trump plan: 20 million vaccines in January’21, 30 in February’21, 50 million monthly thereafter<br>Biden plan: 1 million vaccines every day |
| Non-pharmaceutical interventions | 6 scenarios, including a no-intervention case. The dataset in **Table 4** explores the scenarios. *(For a complete description of the six scenarios, see Table 2 in Li & Giabbanelli <sup>[12]</sup>)* |
| % of population seeking vaccines | From 20% to 60%. Intermediate values are automatically determined to maximize information. |
| Vaccine efficacy | From 88% to 99%. Intermediate values are automatically determined to maximize information. |

**Table 4:** Parameters and values explored by Li and Giabbanelli in the absence of vaccines. When vaccines are present, only 6 combinations to provide broadly different scenarios in **Table 3**.

| Parameter | Values Used |
| --- | --- |
| Mask wearing | In all networks, work and school only, community only |
| Reduction in social contacts | At work and in schools: 5%, 30%. In the community: 10%, 30% |
| When to test quarantined individuals | Start of quarantine, end of quarantine, both |
| Test sensitivity | 55%, 100% |
| Delay in contact tracing | None, 1 week |
| % of contacts of a positive case that can be traced | 20%, 100% |
| When to start contact tracing for an exposed individual | Immediately, upon confirmation from a positive case |

#### 3.1.2 Shamil et al. Model ^[24]^

For the Shamil model, we ran the New York city configuration with 4 different values for 2 parameters using a factorial design, where every possible combination is used. We also performed 10 replications on each combination. The parameters we used are explained in **Table 5**. The 85% smartphone ownership level is based on a survey by the Pew Research Center from February 8, 2021 ^[93]^. The 14-day quarantine is based on the recommendations of the CDC at a time close to when the model was developed, in December 2020 ^[94]^. In total, we generated *n* = 240 data points from this model.

**Table 5:** Parameters and values used in the Shamil *et al*. model dataset.

| Parameter | Values Used |
| --- | --- |
| Days required for contact tracing | 1, 3, 5, 7 |
| Percentage of contacts that can be traced | 0.5, 0.6, 0.7, 0.8 |
| Percentage of the population who own a smartphone | 0.85 |
| Days required in quarantine | 14 |

#### 3.1.3 Silva et al. Model ^[29]^

For the Silva model, we ran the model with a factorial design on 5 different parameters, running each for 60 days in the model. We also performed 30 replications on each combination. The parameters we used and their values are listed in **Table 6**. To verify that we used this model correctly, we replicated *Figure 5a* from the original paper ^[29]^ that used tracked agent states over 60 days with no interventions and a population and grid size of 300. In total, we generated *n* = 13, 500 data points from this model.

**Table 6:** Parameters and values used in the Silva *et al*. model dataset.

| Parameter | Values Used |
| --- | --- |
| Population size | 500, 750, 1000 |
| Grid size | 500, 750, 1000 |
| Type of lockdown | **none, total, half, conditional, vertical** (explanations in **Table 7**) |
| Masks required? | **True, False** |
| Contagion distance | 0.05, 0.25, 0.5, 0.75, 1 |

**Table 7:** Explanations of values for the lockdown type parameter (c.f. **Table 6**).

| Lockdown Type | Description |
| --- | --- |
| **none** | No lockdown |
| **total** | Lockdown required for everyone |
| **half** | Lockdown required for random 50% of the population |
| **conditional** | Lockdown required for anyone in a contagious environment |
| **vertical** | Lockdown required for anyone in an at-risk category or in a contagious environment |

#### 3.1.4 Badham et al. Model ^[30]^

For the Badham model, we ran the model with a factorial design on 4 different parameters, performing 30 replications. All other settings are set to the default values for the model’s 1.1 version. The parameters we altered are explained in **Table 8**. To verify that we used this model correctly, we replicated *Figure 4* from the original paper ^[30]^ that tracked daily hospital admissions per day. In total, we generated *n* = 2, 430 data points from this model.

**Table 8:**
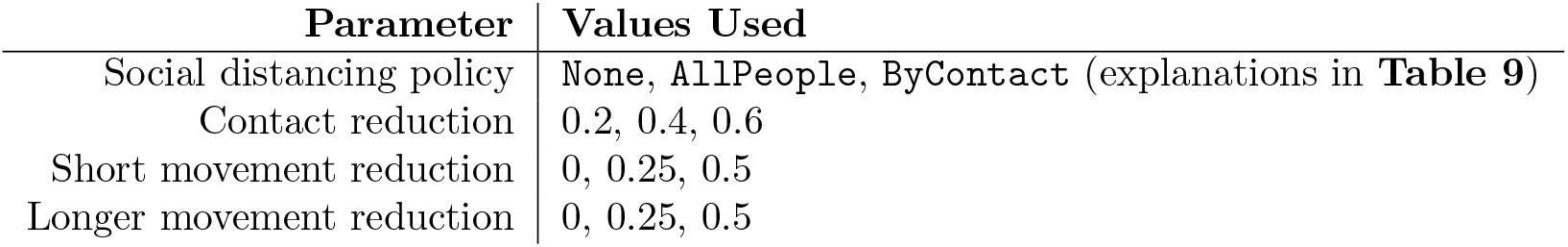
Parameters and values used in the Badham *et al*. model dataset.

**Table 9:** Explanations of values for the social distancing policy parameter (c.f. **Table 8**).

| Distancing Policy | Description |
| --- | --- |
| None | No distancing |
| AllPeople | Everyone reduces their contacts by the “contact reduction” amount |
| ByContact | Probability of contact is reduced by the “contact reduction” amount |

**Figure 4:**
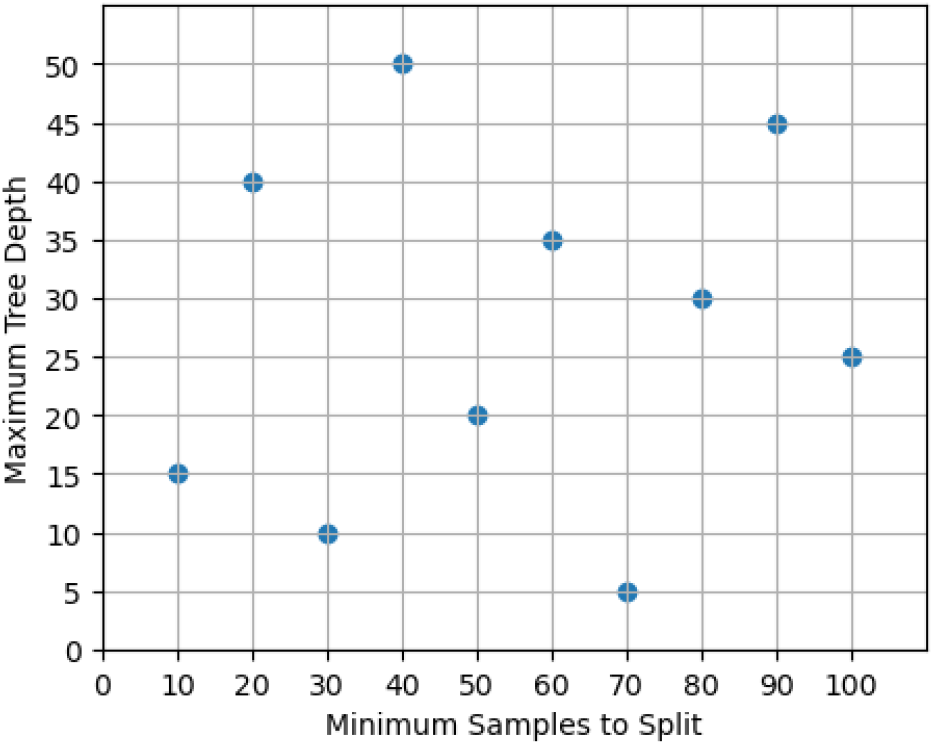
Latin hypercube for `TreeLearner` hyper-parameters.

### 3.2 Normalization & Model Training

To perform our regressions, we used `TreeLearner` regressors from the Orange data mining library (`orange3` version 3.26.0) running on Python (version 3.8.3). When we loaded our datasets, we converted their cumulative infection numbers to proportions of the total population, so that the results from the four different models are comparable. Then, we used a 10-fold cross-validation method to actually train our meta-models. Cross-validation involves separating data into 10 different “folds.” One by one, these folds are used as testing data for our model, while the other nine folds are used to train the model. We also needed to optimize the parameters of our regressor, so we used *hyper-parameter optimization* to maximize our meta-model accuracy.

Hyper-parameter optimization ensures that the regressor is using the most accurate set of model parameters, and it does this by performing a second set of cross-validations, which is known as a *nested cross-fold validation* design. The training data from the first set of folds is divided into a further 10 folds, where one fold at a time is used as a validation set. Then, the remaining nine folds are used to train a model using the different combinations of parameters to select the best combination. To select our parameters, we used a Latin hypercube design, which ensures that the sample space for parameters is covered effectively. We optimized the two parameters that have the most impact on the decision tree regressor:

- the *maximum depth* of the tree. A smaller depth forces the meta-model to be simplified. We considered values from 5 to 50 by steps of 5.
- the *minimum number of samples* to split a node. The machine learning algorithm stops sooner and delivers a simpler model if we raise the minimum number of samples. We examined values from 10 to 100 by steps of 10.

For details on the impact of these parameters on a decision tree, we refer the reader to two recent examples of optimization ^[95, 96]^. The optimization process was conducted using a Latin hypercube with 10 different samples (**Figure 4**).

### 3.3 Model Evaluation

Once the hyper-parameter optimization selected the most accurate set of parameters, these parameters were used to build the model for the outer folds. Then, we were able to analyze how the accuracies of the models changed in comparison with the amount of data that was used to train them by calculating the root-mean-square error for their predicted infected proportions (**Equation 1**). RMSE is a common error metric for regression models ^[97, 98, 99]^ and it is useful because the units for RMSE are the same as the units for the simulation output (i.e. our errors for this paper are measured in proportions of the population).

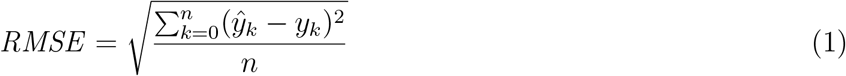

## 4 Results

The ABMs used for this study were chosen as we could access their code and verify our use of the model vis-a-vis published figures in peer-reviewed manuscripts. The first subsection focuses on this verification effort, by contrasting the results that we obtained from the model with the authors’ published results; figures were all reproduced with the authors’ permission. The second subsection is centered on the machine learning results. To provide full transparency and support replicability efforts regarding machine learning, the training data used in this subsection is *openly accessible* on the third-party repository https://osf.io/d7vqa/, provided by the Open Science Framework.

### 4.1 Agent-Based Model Verification

As noted in section 3.1.1, the model by Li and Giabbanelli ^[12]^ was not subject to verification, since we directly used data produced by the model. That is, we already had access to its full simulation results. In contrast, the other two models had to be verified since we did not have a spreadsheet of results to use and hence we ran the code provided by the authors. The verification sought to replicate the published work of the authors, to ensure that we were using their model adequately.

For the Shamil model, we replicated *Figure 5a* from Shamil, et al. ^[24]^ at six different contact tracing levels (0, 0.6, 0.7, 0.75, 0.8, and 0.9). Qualitatively, we note that shapes and trajectories of the infections are similar in our replication (**Figure 5**). However, values occasionally differ and some of the curves for the higher trace levels also overlapped in our replication, which is not something found in the original. Based on previous attempts at replicating simulation models ^[100]^, a possibility is that the high variability in the model’s output results in noticeable differences across individual runs and only a large number of runs (e.g., to achieve a 95% confidence interval) would allow to create comparable curves. In other words, the discrepancies are a likely consequence of output variability in the published work.

For the Silva model, we replicated *Figure 5a* from Silva, et al. ^[29]^ using a population size and grid size of 300 (**Figure 6**). The state counts in our replication closely follow the average state counts in the original figure.

**Figure 5:**
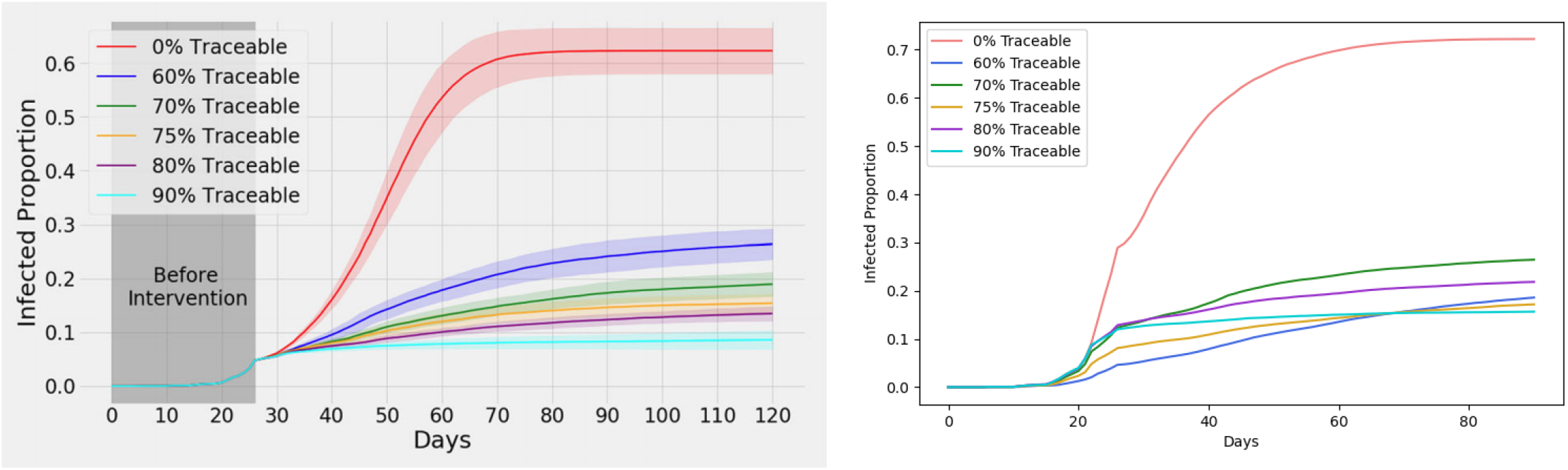
*Figure 5a* from Shamil, et al. ^[24]^ (left) with our replication (right) using 5 repetitions. *The left figure is reproduced with permission from the authors*.

**Figure 6:**
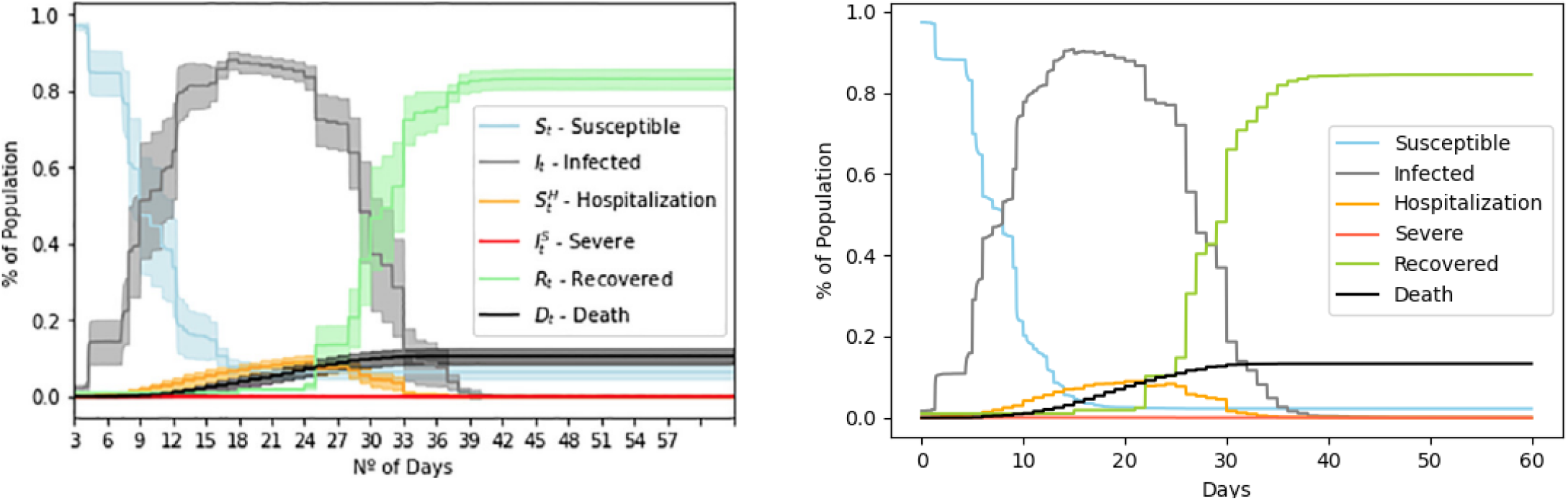
*Figure 5a* from Silva, et al. ^[29]^ (left) with our replication (right) using 10 repetitions. *The left figure is reproduced with permission from the authors*.

For the Badham model, we replicated *Figure 4* from Badham, et al. ^[30]^ using the model’s `atJul13` scenario (**Figure 12**). The shape and values in our replication closely follow the original figure.

### 4.2 Machine Learning Results

The root-mean-square error for the Li and Giabbanelli model ^[12]^ *without vaccines* is shown in **Figure 8**. The RMSE is around 0.0795 *regardless of the sample size*, which shows that running as little as 5% of the experiments from the ABM suffices to then infer the rest via machine learning. However, we note that the error margins do benefit from an early increase in data, of up to 30% of the sample size. The situation changes when vaccines are introduced. For both the Trump administration’s vaccine plan and the Biden administration’s plan, we see that the RMSE strictly decreases as the sample size increases (**Figure ??**). The error can be as low as 0.0034, while noting that the decrease is essentially from a low initial error (RMSE of about 0.0041 to 0.0043 at 5% of the sample size) to a slightly lower error.

**Figure 7:**
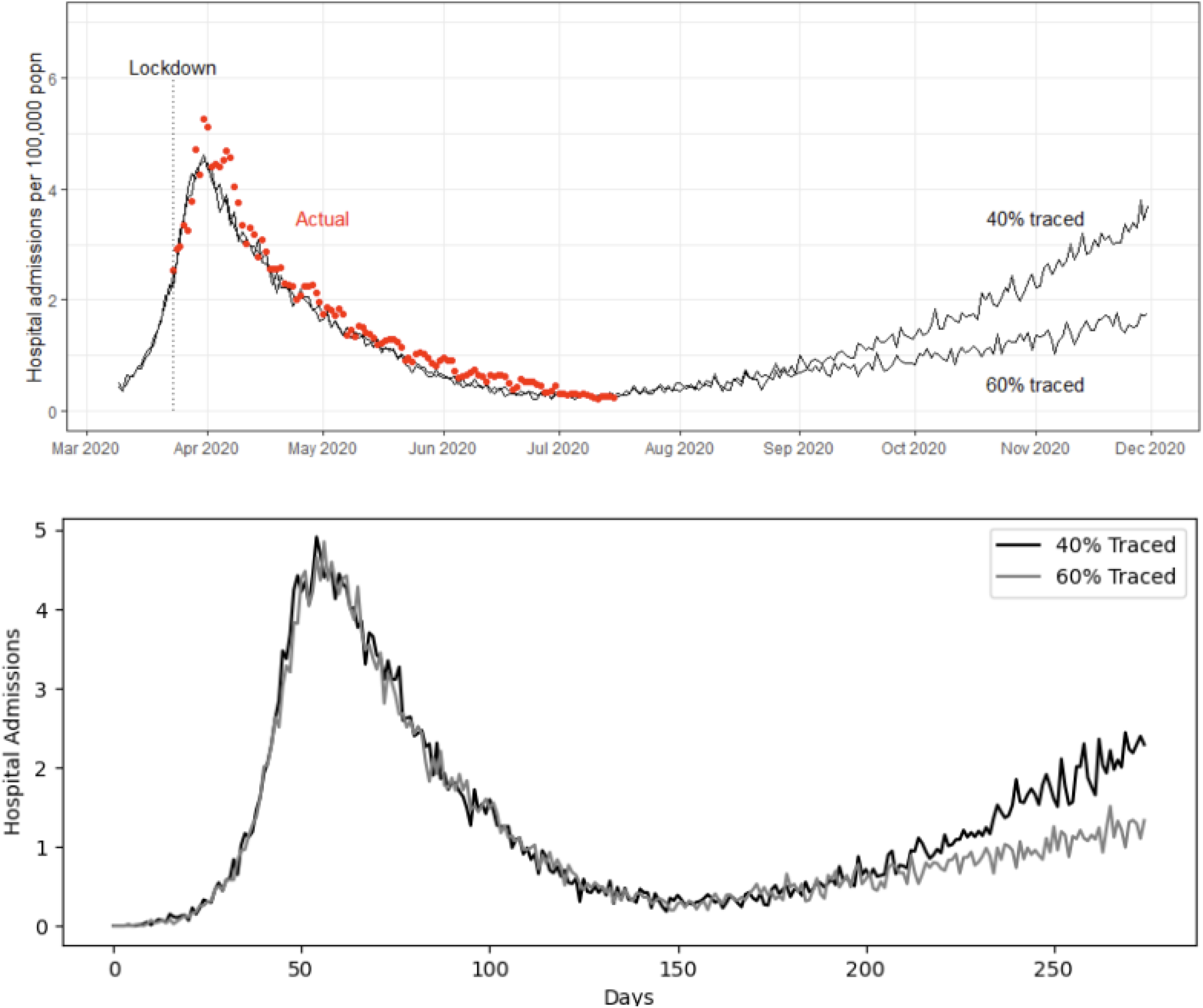
*Figure 4* from Badham, et al. ^[30]^ (top) with our replication (bottom) using 1000 repetitions. *The left figure is reproduced with permission from the authors*.

**Figure 8:**
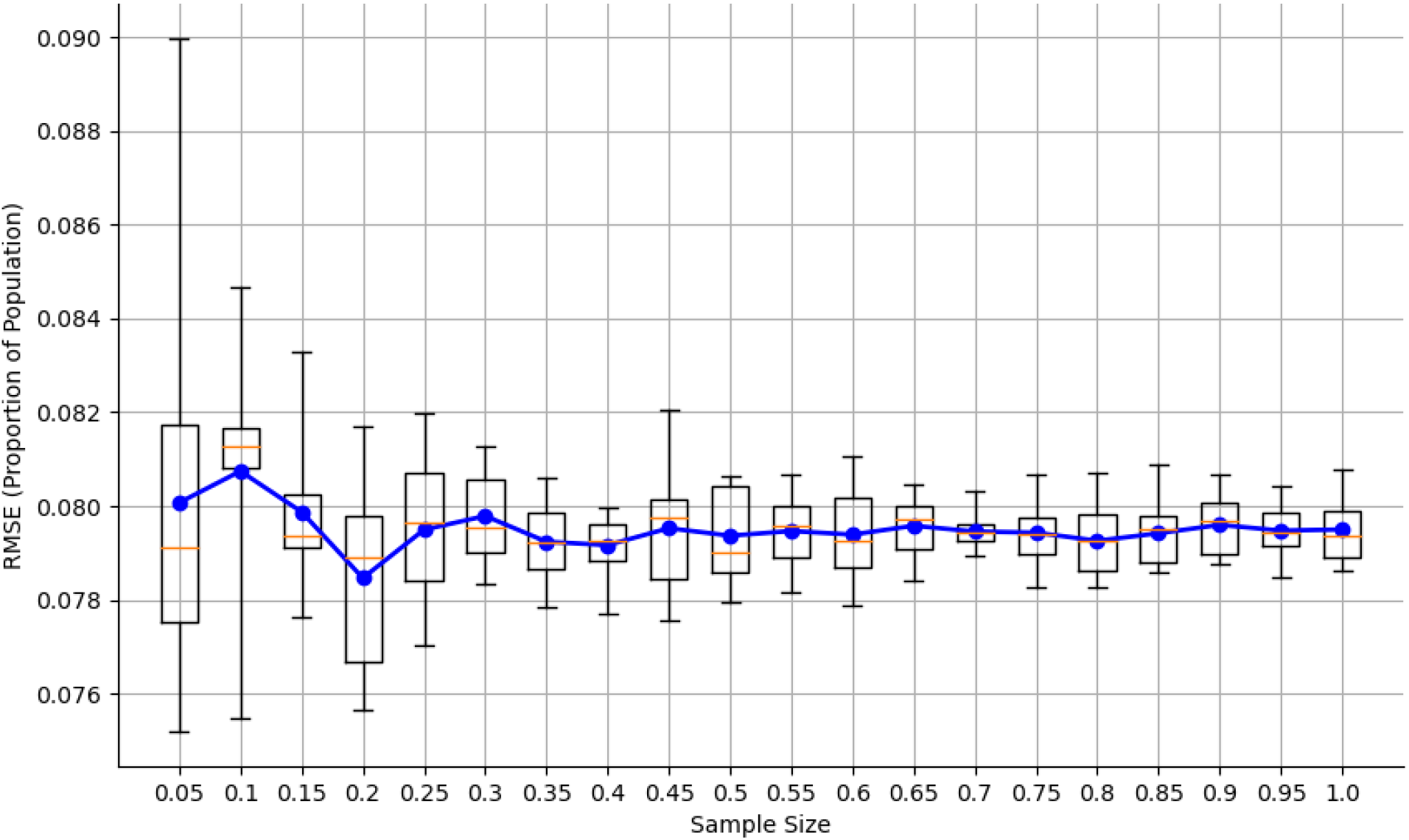
Graph of the root-mean-square error of the predicted proportion of the population that was infected at different sample sizes for the Li and Giabbanelli dataset without vaccines.

**Figure 9:**
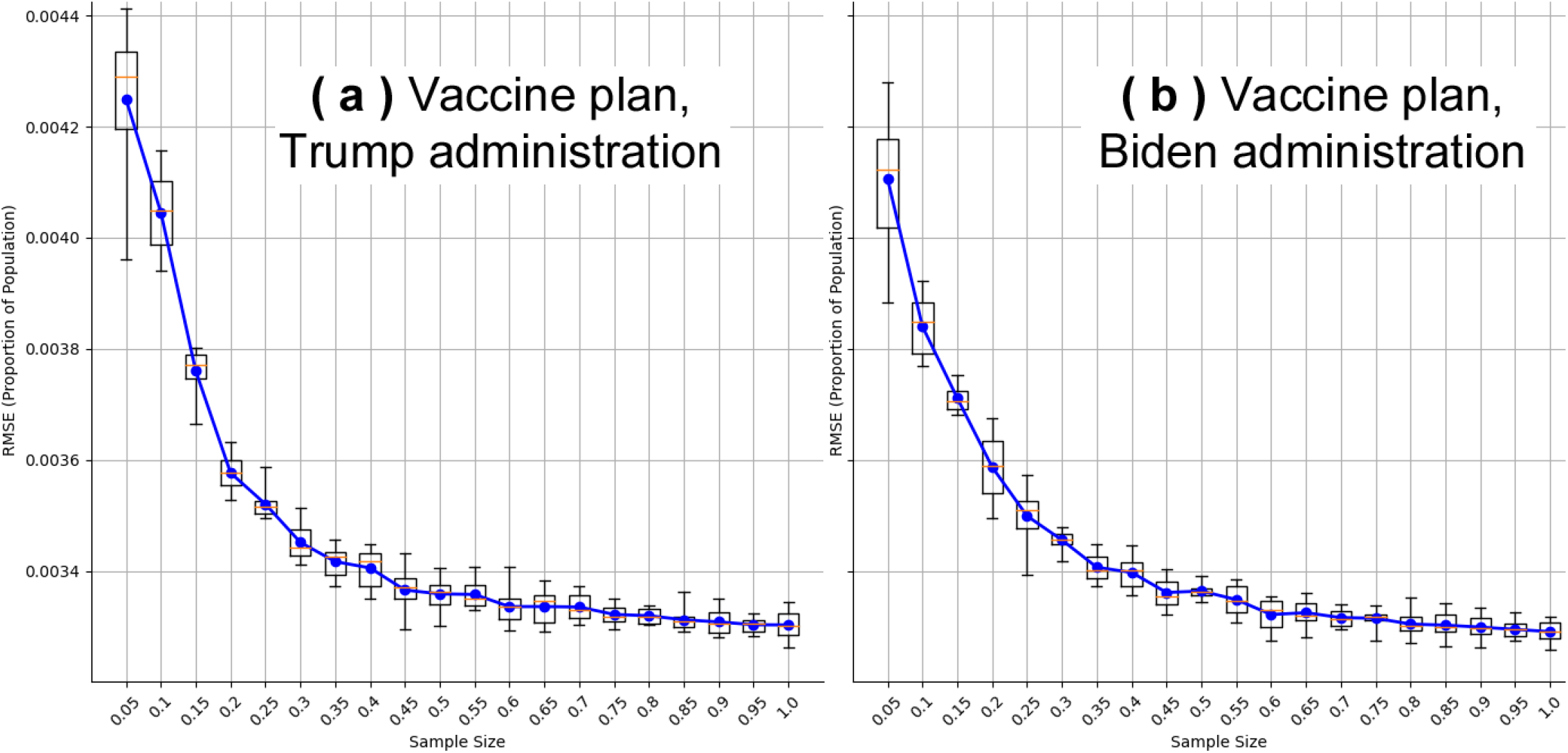
Graph of the root-mean-square error of the predicted proportion of the population that was infected at different sample sizes for the Li and Giabbanelli dataset using the Trump administration vaccine plan (a; left) and Biden administration vaccine plan (b; right).

Different situations are observed among the other three models. For the Shamil *et al*. model ^[24]^, results show an error around 0.065 when using most of the data (**Figure 10**). Most interestingly, the error has a non-monotonic relation with the sample size: it is lowest at the smallest sample size (0.1%) and at about half of the sample (0.45%), but higher in between. For the model by Silva *et al*. ^[29]^, we see a decrease in RMSE as th sample size increases. Again, we note that the gains should be contextualize given the small range of the RMSE: 10% of the dataset suffices to achieve and error of 0.028 and even taking the full dataset only brings it down to 0.0225 at the best. Finally, the model by Badham *et al*. ^[30]^ shows a decrease of RMSE as the sample size increases up to 50% of the dataset, and then the error plateaus. Relative to its scale, we emphasize that the total decrease in RMSE between the 5% and 100% sample sizes is only around 0.01.

**Figure 10:**
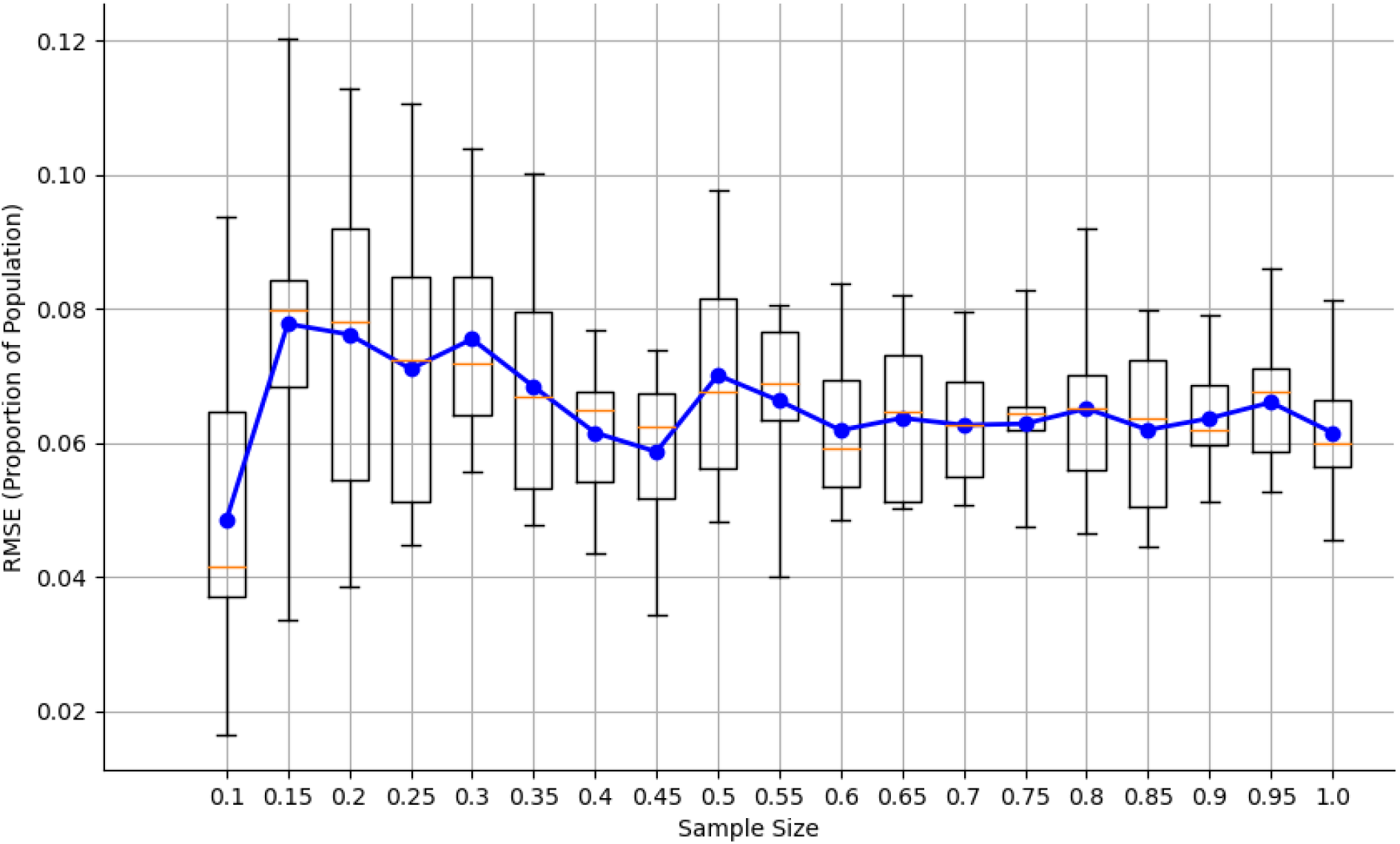
Graph of the root-mean-square error of the predicted proportion of the population that was infected at different sample sizes for the Shamil, et al. dataset.

**Figure 11:**
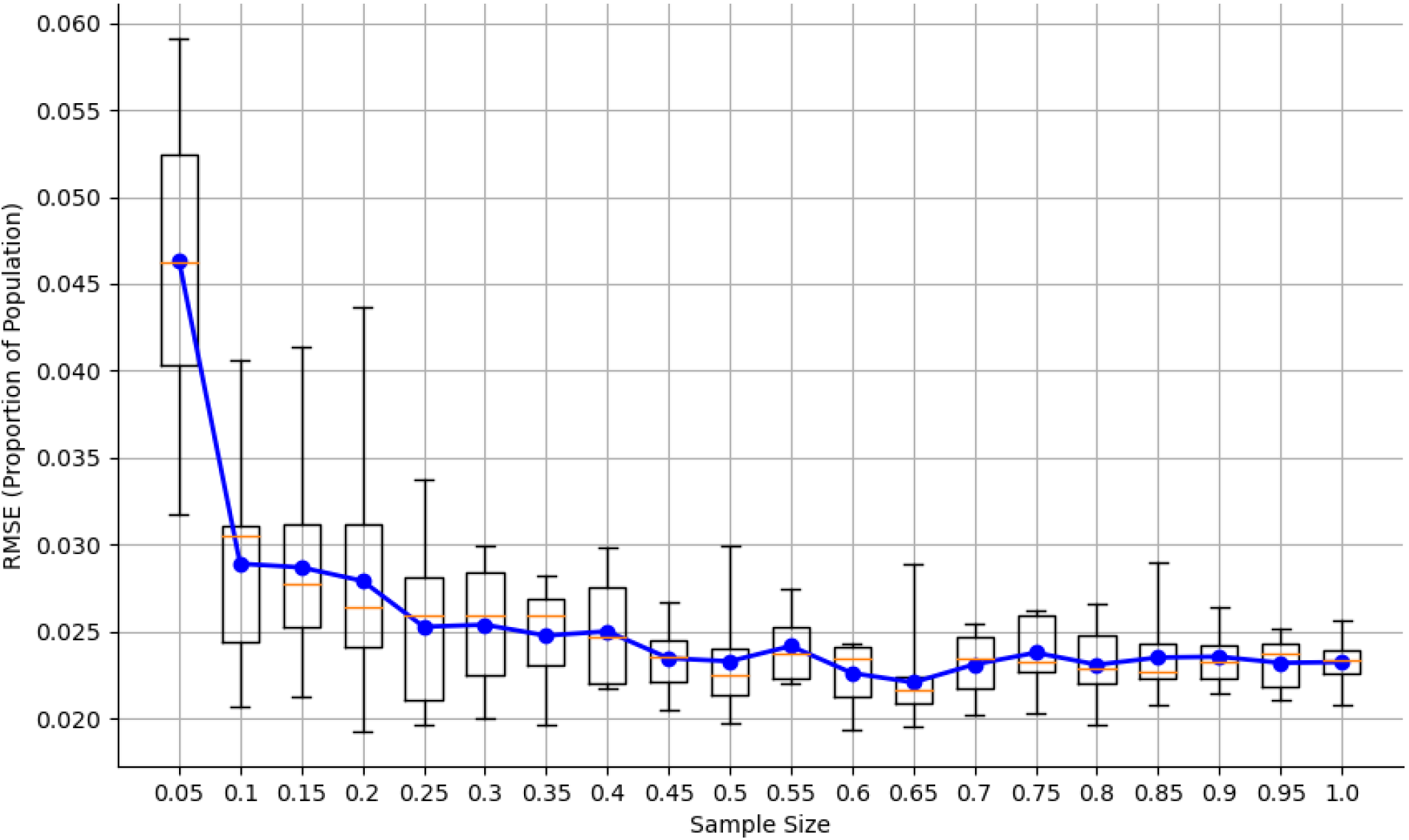
Graph of the root-mean-square error of the predicted proportion of the population that was infected at different sample sizes for the Silva, et al. dataset.

**Figure 12:**
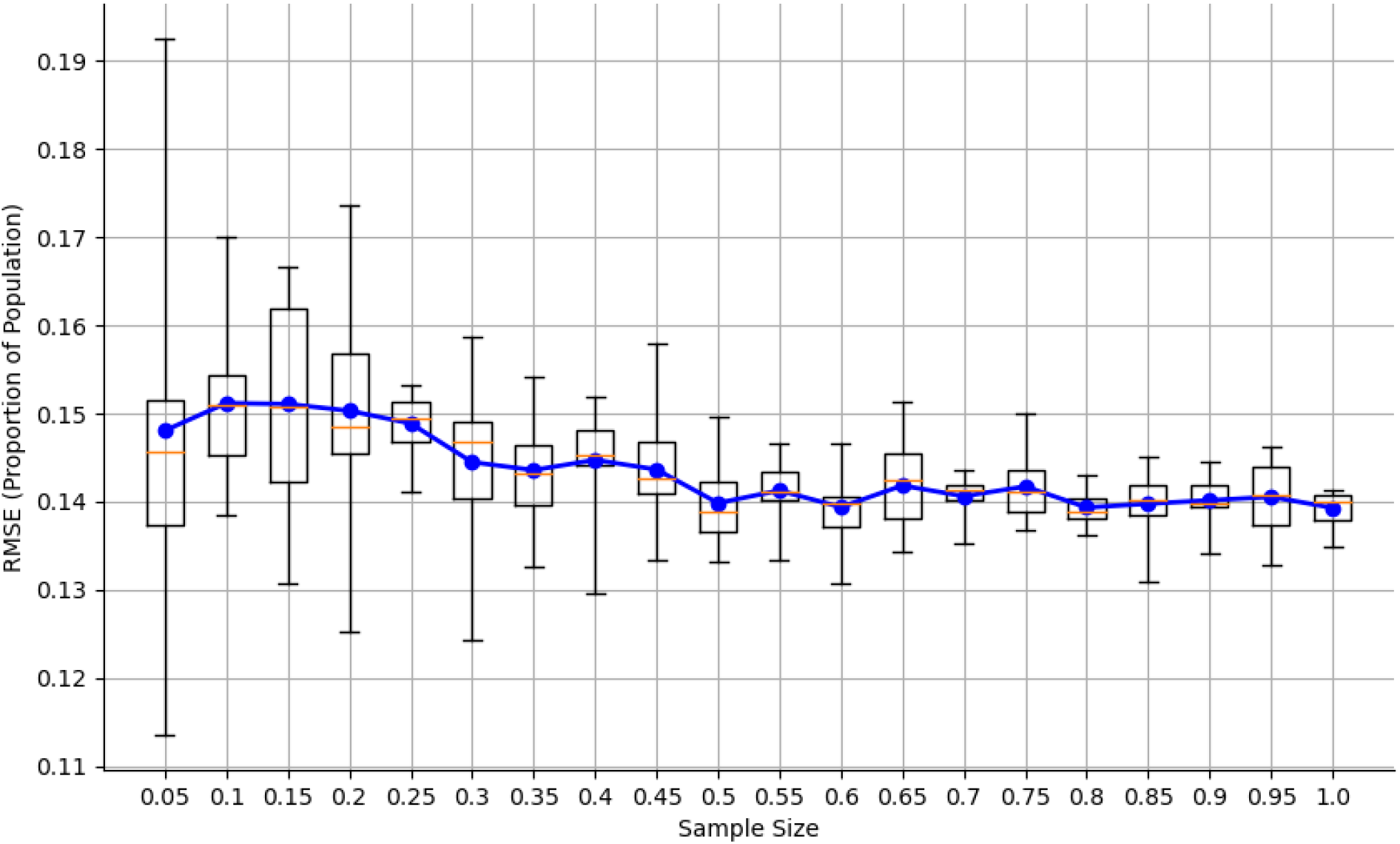
Graph of the root-mean-square error of the predicted proportion of the population that was infected at different sample sizes for the Badham, et al. dataset.

## 5 Discussion

While machine learning and simulations have both been independently used to model COVID-19 ^[65]^, our study is the first to combine these techniques to examine whether access to COVID-19 Agent Based Models could be democratized by making them more computationally affordable for end users. Specifically, we analyzed how the amount of data used to train a regression meta-model affects its accuracy and differentiated situations where more data is required from situations where small amounts of data are sufficient. This analysis contributes to the broader literature on assessing and simplifying COVID-19 models, which has already established that the number of parameters could be decreased significantly ^[59]^. Our results shows that models which do not have strong interventions like lockdowns or vaccines do not require as much training data, hence it is possible to run *few* computationally expensive simulations and then switch to an inexpensive metamodel. The Covasim model with no vaccines and the Shamil model showed no discernible downward trend in their root-mean-square error values, so adding more training data would not guarantee an increase in model accuracy. While the Badham model showed a slight decrease in RMSE, it only decreased by 0.01 from 5% to 100% sample size. Conversely, the models which did have strong interventions showed a strong downward trend in their RMSE values. Both Covasim models that used vaccines only began to stabilize after 60% of the data was used in training, and while the Silva model stabilized slightly faster, it took around 45% of the data to do so. Note that the RMSE values were low to start with (e.g., less than 0.0045 for Covasim with vaccines), so the initial error may already be tolerable for some end users as part of a tradeoff between accuracy and computational needs. There are three main limitations to our work. First, we focused on peer-reviewed COVID-19 ABMs that we could run to obtain data and for which we could verify our use of the model vis-a-vis published results. As shown in previous calls for transparency in COVID-19 modeling, modelers do not systematically provide their code ^[101]^. In a transparency assessment, Jalali and colleagues found that most models do not share their code ^[102]^, which echoes similar observations about practices in Agent-Based Modeling across application domains ^[103, 104]^. Our criteria thus meant that we could only assess a subset of existing models and it is possibly that different trends or initial error levels are observed in other models. Second, several key parameters and assumptions regarding COVID-19 continue to change. For example, a study on almost 100,000 volunteers in July 2021 found a vaccine effectiveness of 49%, which is less than the *lowest* value assumed in some of the previous modeling studies ^[12, 105]^. Other studies have shown that vaccinated individuals have a comparable viral load to unvaccinated ones within the first few days ^[106, 107]^, whereas the flow diagrams used in many models have considered that vaccinated individuals were fully removed from the population. Our conclusions are limited to COVID-19 ABMs developed so far, since future models may exhibit markedly different dynamics. In particular, bifurcation may be present in future models, which would require an analysis of models by clusters of trajectories rather than average dynamics ^[108]^. Finally, the size of the datasets that we generated were limited by the high computationally needs of the COVID-19 ABMs. For example, the Shamil model required around 8 hours per run on a node of a high-performance computing cluster. Given this limitation, our conclusions focus on the *trend* (e.g., is there a reduction in error as more simulations are used?) rather than on a precise point-estimate of the error (e.g., exactly how many simulations are required to achieve a given error level).

There are several potential avenues for future work. We focused on predicting the *final* result of the simulation, but there would also be merit to predicting the *time series* of outputs. Although the use of meta-modeling of simulations for time series is more common for financial simulations ^[109]^, there could also be an opportunity for future studies to apply these techniques in the case of COVID-19 ABMs. In particular, several such ABMs (e.g., COVID-Town ^[110]^) have been developed to perform a joint analysis of economic and epidemic dynamics, hence they are particularly interested in the *shape* ^[111]^ of the economic recovery over time (i.e., the time series).

Another possibility would be to explore the prediction of *multiple outputs*. Indeed, various stakeholders may be interested in the effects of a COVID-19 intervention on different outcomes ranging from epidemiological (e.g., number of new cases) to logistical (e.g., spare capacity in intensive care units) and economical (e.g., loss in productivity or revenues). While we may expect that *more* simulations are necessary to train an accurate metamodel able to predict several partially correlated outputs, the specific *relationship* is still a question. That is, we still need to establish how the number of predicted outputs raises the need for simulation data.

## 6 Conclusion

In this paper, we analyzed how the amount of data used to train a simulation meta-model affected the accuracy of the model. We found that for models with no strong interventions such as vaccines or lockdowns, a small amount of data could generate a model with similar accuracy to one trained on a much larger amount of data. However, models which had strong interventions took large amounts of data to train a model that achieved a stable accuracy. These results indicate that modeling the spread of COVID-19 without strong interventions can be done with very little data, but when stronger interventions are considered, much more data is required to train an accurate model.

## Data Availability

Data is publicly available on a third-party repository (Open Science Framework) at https://osf.io/d7vqa/ under "COVID-19 Metamodeling - Training Datasets"

## Acknowledgments

For each of the three models used in this paper, we thank the corresponding authors (Dr Mohammad Sohel Rahman, Prof. Dr. Petrônio Cândido de Lima e Silva, Dr. Jennifer Badham) for giving us permission to reuse their images in our verification process. We also appreciate the assistance of Dr. Badham in operating the model.

## Conflicts of Interest

The authors declare that they have no conflicts of interest.

## Notes

### Competing Interest Statement

The authors have declared no competing interest.

### Funding Statement

The authors received no financial support for the research or authorship of this manuscript.

### Author Declarations

Not applicable (no human data used).

